# Risk Factors and Outcomes of Pulmonary Hemorrhage in Preterm Infants born before 32 weeks

**DOI:** 10.1101/2024.06.22.24309343

**Authors:** Gieng Thi My Tran, Nhat Phan Minh Nguyen, Nguyen Phuoc Long, Duc Ninh Nguyen, Thu-Tinh Nguyen

**Author notes:** **Corresponding Author:** Tinh-Thu Nguyen, MD, PhD; Department of Pediatrics, University of Medicine and Pharmacy at Ho Chi Minh City, 217 Hong Bang Street, District 5, Ho Chi Minh City, 700000, Vietnam.

## Abstract

**Background:** Pulmonary hemorrhage (PH) is a catastrophic event associated with significant morbidity and mortality among preterm infants. Understanding PH risk factors in preterm newborns, particularly those in low-to-middle-income countries like Vietnam, remains limited. This study aimed to investigate the risk factors and short-term outcomes of PH in very preterm infants.

**Methods:** We conducted an observational study of newborns aged < 72 hours with gestational age < 32 weeks, admitted to our unit from April 1, 2018 to March 31, 2019.

**Results:** Of 118 recruited newborns, 28 (23.7%) had PH. The logistic regression analysis showed that intubation within 24 first hours, blood transfusion, and coagulation disorders within the first 3 days were risk factors for PH (aOR = 4.594, 95% CI = 1.200-17.593; aOR = 5.394, 95% CI = 1.243-23.395 and aOR = 7.242 95% CI = 1.838-28.542, respectively). Intra-ventricular hemorrhage (IVH) and mortality rates were higher in patients with PH compared to those without (p<0.001). The length of invasive ventilation was longer in the PH group (p<0.001).

**Conclusion:** We have identified intubation, blood transfusion, and coagulation disorders shortly after birth as risk factors for PH in very preterm infants, which was associated with increased mortality and occurrence of IVH.

**Impact:**

1. High incidence and mortality of pulmonary hemorrhage in preterm infants < 32 weeks’ gestational age with respiratory distress in a Neonatal Intensive Care Unit in Vietnam.
2. Pulmonary hemorrhage should be considered in the clinical deterioration of preterm infants given invasive ventilation in the first 2-4 days of life.
3. Independent risk factors for pulmonary hemorrhage: intubation in the first 24 hours, coagulation disorders, and transfusion of blood products in the first 3 days of life.
4. Urgent need to seek diagnostic criteria for intraventricular hemorrhage as soon as pulmonary hemorrhage occurs.

## Introduction

Pulmonary hemorrhage (PH) is a life-threatening condition with blood extravasation into the alveoli, causing significant morbidity and mortality in hospitalized preterm infants. It occurs in up to 1.2% of total live births^1^, with a particularly high incidence in preterm infants (up to 5%)^1^. PH currently has no preventive treatment and is most commonly diagnosed during the first few days of life; its mortality rates could rise to 50%, especially in extremely preterm infants^2^. Its diagnostic criteria include the aspiration of secretions from the trachea or endotracheal tube^3,4^. The severity can vary from a mild, self-limited disorder to a massive, deteriorating, and end-stage syndrome. Upon PH diagnosis in preterm infants, invasive ventilation and prolonged intensive care are often required, further elevating the risks of other significant morbidity and mortality^3,43^. In preterm infants, it is often manifested from pulmonary edema and hemodynamically significant patent ductus arteriosus (hsPDA)^3,4^. Asphyxia, prematurity, intrauterine growth restriction, early-onset sepsis, patent ductus arteriosus, and surfactant replacement have been shown as major risk factors of PH in several geographical settings^2–5^. Awareness of PH risk factors may help clinicians realize the early symptoms and develop prompt strategies for PH management.

The already reported risk factors of PH in preterm newborns are variable across different settings. In Vietnam, PH incidence and associated factors are unknown. Therefore, this study aimed to explore the perinatal associated factors and short-term outcomes of PH in very preterm newborns in a neonatal intensive care unit (NICU) in Vietnam, which will provide future benefits for early triage and the management of critically ill patients.

## Materials and Methods

### Study design

This retrospective observational study was conducted at the NICU of the Children’s Hospital 1 (a level IV NICU) from April 1^st^, 2018 to March 31^st^, 2019 in Ho Chi Minh City, Vietnam. The study protocol was reviewed and approved by the Institutional Review Board of Children’s Hospital 1 (IORG0007285, FWA00009748, Approval number: 2784/QĐ-BVNĐ1, On October 16^th^, 2019). Written informed consent was waived by the Institutional Review Board because this was a retrospective study. There was no patient contact, and some of the patients were deceased.

The inclusion criteria include: (i) gestational age at birth < 32 weeks, (ii) being admitted to the NICU within 72 hours following the birth, and (iii) absence of congenital anomalies, including congenital heart disease other than patent ductus arteriosus (PDA), structural anomalies of the gastrointestinal tract, and other anomalies requiring surgery. Patients developing PH prior to admission to our center were also excluded.

PH diagnosis was performed according to the international standard with hemorrhagic secretions aspirated from the trachea or endotracheal tube, accompanied by clinical deterioration, which included the requirement of intubation and ventilation or the need for an increased fraction of inspired oxygen (FiO2) by at least 10% ^3^. A chest X-ray was also taken to confirm diffuse pulmonary infiltrates.

Information during the pregnancy and hospitalization was collected retrospectively from the medical notes. Maternal history including antenatal corticosteroids, complications of this pregnancy (gestational diabetes, pre-eclampsia/ eclampsia, hypertension, urinary infection), intrapartum fever. Neonatal baseline details including gender, gestational age (calculated from the first day of the last menstrual period or prenatal obstetric ultrasonography in the first trimester), birth weight, mode of delivery, small for gestational age (SGA < 10 percentile on Fenton chart), intubation within first 24h of age, treatment with surfactant, multiple doses of surfactant prior the onset of PH, coagulation problems within the first 3 days, blood transfusion within the first 3 days (prior the occurrence of PH), platelet count < 100000/mm3, hsPDA. Echocardiography was used to diagnose PDA by a cardiologist from day 3 – 7 and repeated when needed. The hsPDA was managed by consensus among neonatologists and cardiologists, and oral ibuprofen or intravenous acetaminophen was given to close the PDA. Surgery was performed in cases of contraindications to ibuprofen/ acetaminophen or failed 2 courses of ibuprofen/acetaminophen therapy.

The study’s primary outcomes include the incidence of PH. Secondary outcomes include PH risk factors, the rate of mortality, IVH, retinopathy of prematurity (ROP) and bronchopulmonary dysplasia (BPD), length of invasive ventilation, and NICU stay in patients with vs. without PH. IVH was identified by cranial ultrasound and classified based on Papile criteria. ROP was screened by an ophthalmologist when infants were 4 weeks of age or 31 weeks of postmenstrual age, whichever came last. BPD was defined as the requirement for oxygen supplementation at 36 weeks postmenstrual age or at 28 days old.

#### Statistical analysis

The data were imported into Excel. Descriptive statistical analysis was used to describe the characteristics of preterm newborns and their mothers. The normally distributed results are reported as the mean and standard deviation (SD); the remaining results are reported as the median, interquartile range (IQR), or percentage. Chi-squared test, Mann-Whitney test, Fisher’s test, Student’s *t*-test, and logistic regression model were used for statistical analysis. Multiple logistic regression was applied to classify patients into the binary state of PH (Yes/No). Since the sample size was relatively small (N =118) and the PH class ratio was imbalanced (N_Yes_ = 28, N_No_ = 90), a ℓ_2_-penalization was applied to the logistic regression model to alleviate the potential bias in classification and coefficient estimation^6^. A nested *k*-fold cross-validation procedure was conducted to assess the performance of the classification model. The outer loop (k = 5) is for splitting training (70%) and test (30%) data, while the inner loop (k = 10) is for hyperparameters tuning. The generalization performance of the model was obtained from the nested cross-validation procedure by taking the average for each metric over all the outer-loop folds. The area under the curve (AUC) of the Receiver Operating Characteristic curve was used as a metric to assess model performance. Moreover, the ridge logistic regression was interpreted, and adjusted odds ratios (aORs) with their corresponding 95% confidence intervals (CIs) were computed based on regression coefficients and standard error. Specifically, a ridge logistic regression model was first trained on the whole dataset using a 10-fold cross-validation procedure to select the best-performing hyperparameters. Subsequently, using the optimal set of hyperparameters, we fitted the ℓ_2_-penalized logistic regression model on the whole dataset to estimate the regression coefficients for each clinical covariate. Model training and validation were conducted using the *caret* package (version 6.0-94)^7^ in R version 4.3.2. Before the analyses, categorical variables were dummy encoded, while continuous variables were centered and scaled.

## Results

### Baseline characteristics

From April 1^st^, 2018 to March 31^st^, 2019, there were 171 preterm newborns with gestational age < 32 weeks admitted to the NICU of Children Hospital 1. One had severe congenital heart disease, one had hydrocephalus, five cases had congenital gastrointestinal anomalies, and 46 were admitted after 72 hours of age. There were a total of 118 patients enrolled in the study: 28/118 (23.7%) patients were diagnosed with PH, and 21/28 (75%) patients in the PH group were extremely preterm newborns. The male/ female ratio was 0.96. Other baseline parameters comparing patients with vs. without PH are depicted in Table 1.

**Table 1:**
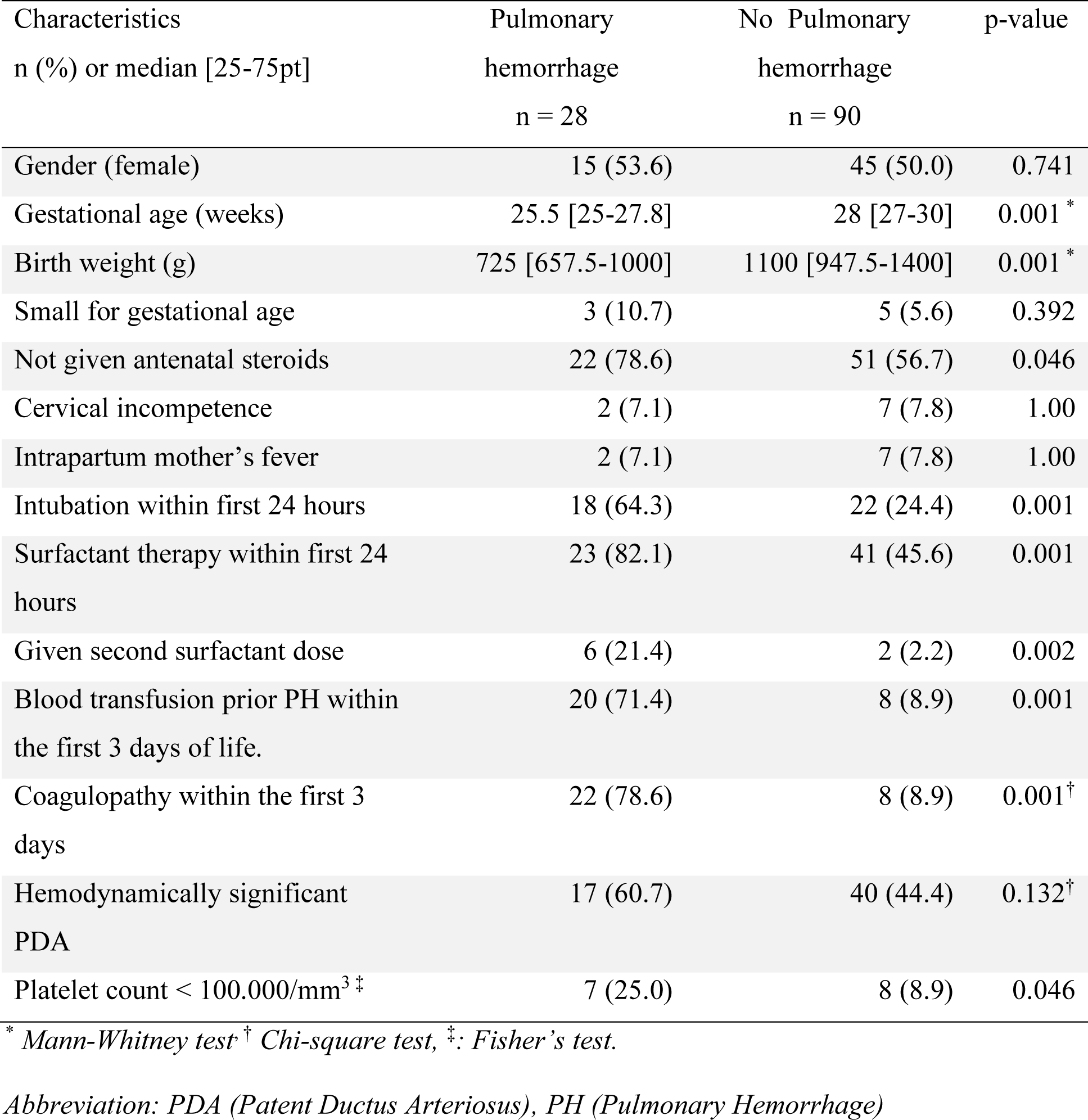
Comparison of clinical characteristics between patients with and without pulmonary hemorrhage.

### Clinical manifestations

The median time for the onset of PH is 47 (32.5-73) hours. The earliest and latest cases in the PH group happened at 28 hours and 144 hours, respectively. It took 38 (26-48.5) hours from the first dose of surfactant to PH manifestation.

The main clinical characteristics of the enrolled patients are also described in Table 1. The analysis of neonatal factors showed that newborns with PH had lower gestational age and birth weight (p < 0.001). Additionally, infants in the PH group were more likely to be intubated in the first 24 hours, treated with surfactant, had been transfused, and had coagulation problems.

Risk factors for PH in this cohort were analyzed via a logistic regression model. The model exhibited considerable performance in classifying patients who underwent PH and those who did not [AUC and standard deviation (SD) from the nested cross-validation = 0.863 ± 0.065] (Figure 1). The aORs of clinical covariates and their corresponding 95% CIs were presented in Figure 2 and Table 2. Three covariates were significantly associated with PH. In detail, patients who had coagulation dysfunction in the first 3 days increased the odds of acquiring PH with an aOR = 7.242 (95% CI = 1.838-28.542). In the same vein, blood transfusion prior to PH in the first 3 days (aOR = 5.394, 95% CI = 1.243-23.395) and intubation in the first 24 hours (aOR = 4.594, 95% CI = 1.200-17.593) were positively associated with undergoing PH.

**Figure 1.**
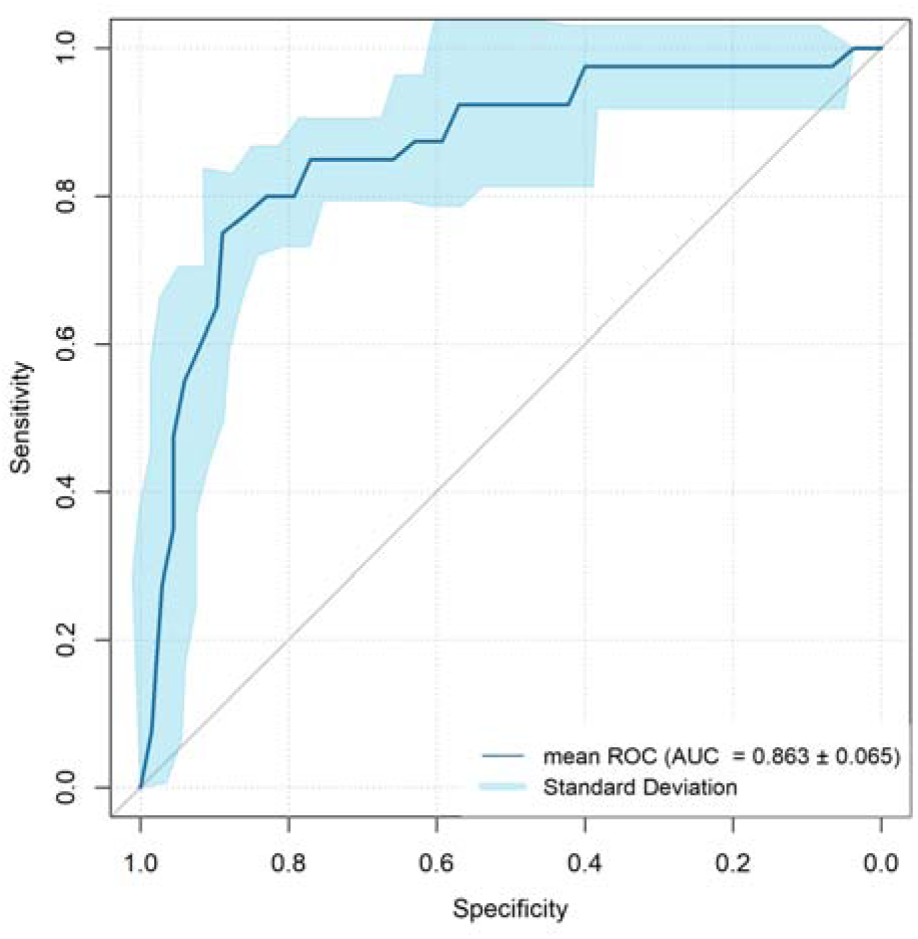
The receiver operating characteristic curve of the ridge logistic regression model classifies patients who acquired PH and those who did not. The total sample size was 118 patients (N_Yes_ = 28, N_No_ = 90).

**Figure 2.**
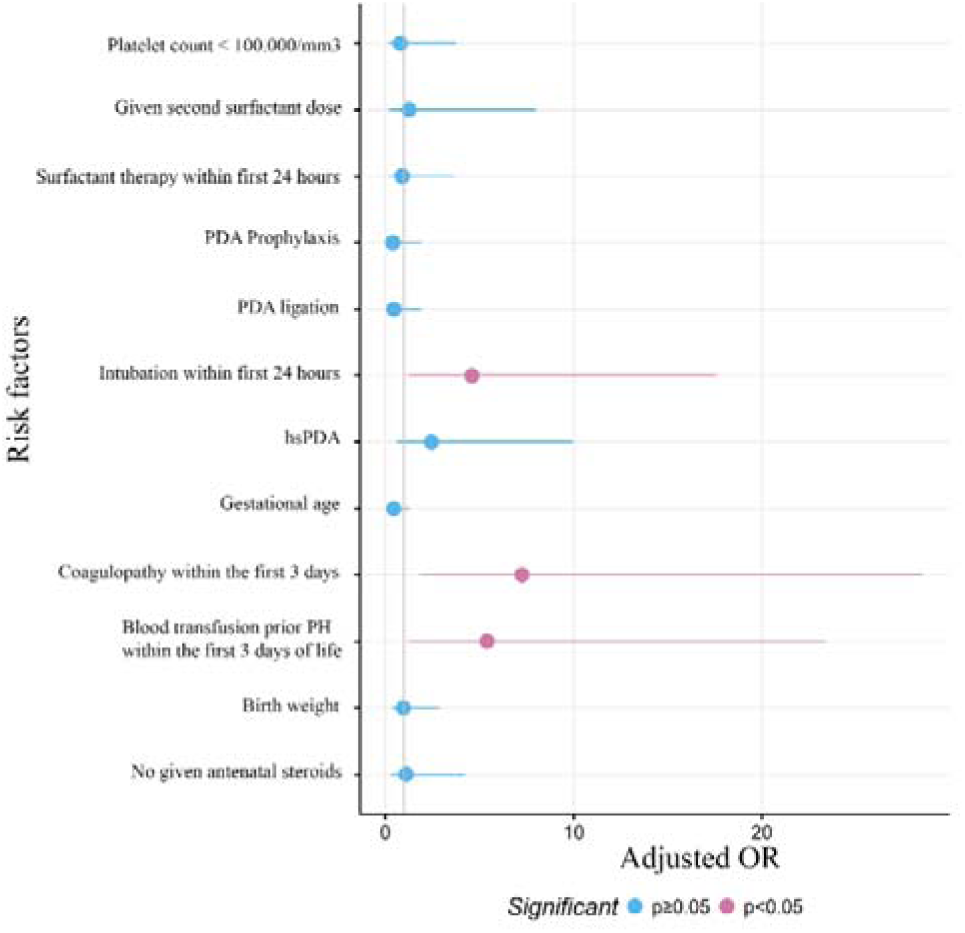
Odds ratio of factors associated with pulmonary hemorrhage. Adjusted odds ratios and corresponding confidence intervals estimated from the ridge logistic regression model. The confidence level is 95%, and non-significant results are presented in blue.

**Table 2:**
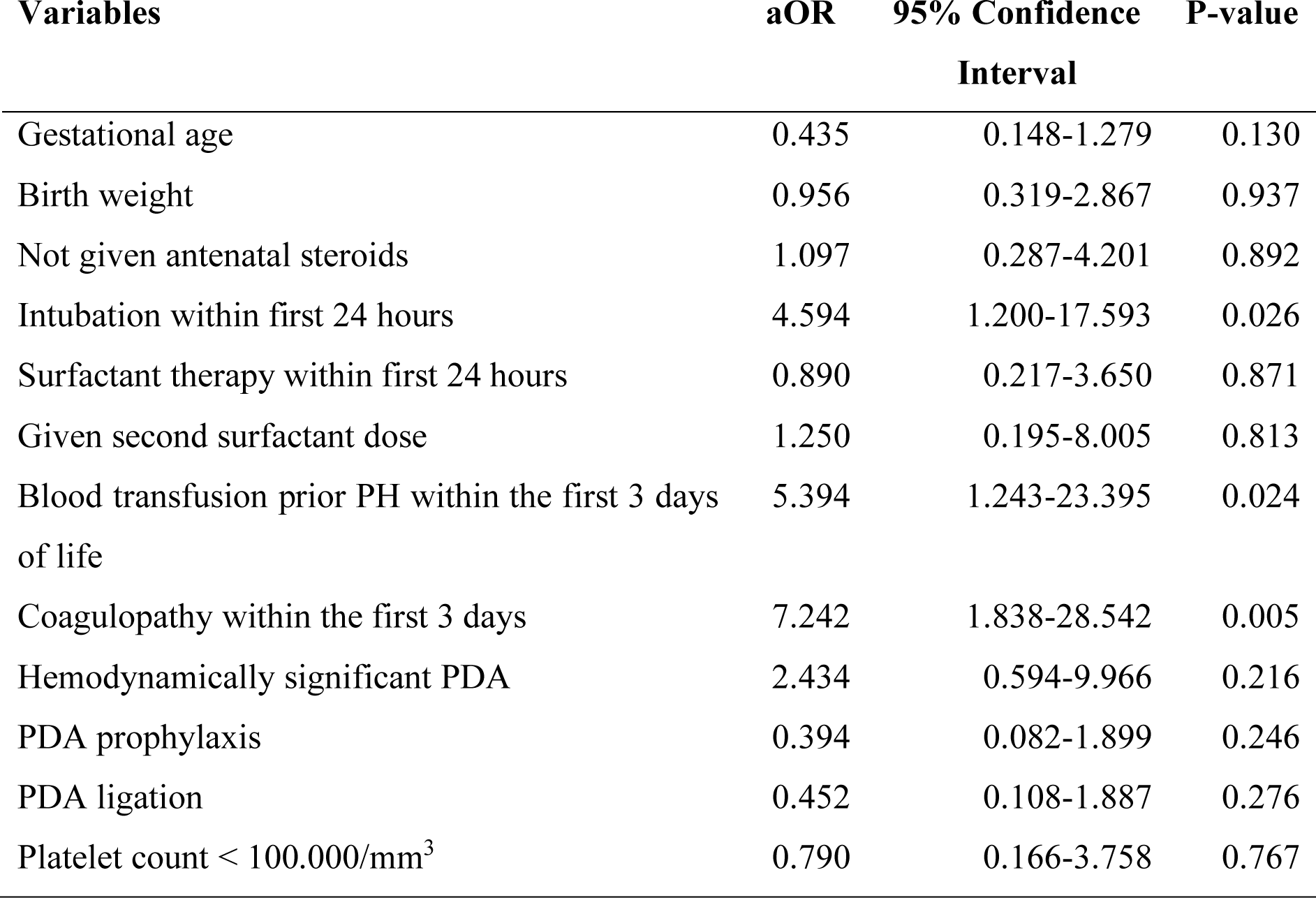
Adjusted odds ratio and 95% confidence intervals estimated from ridge logistic regression model.

Short-term outcomes between PH and no-PH group are described in Table 3. Newborns who had PH showed a higher mortality rate (64.3% vs 25.6%, p= 0.001). Grade II to IV intraventricular hemorrhage was more frequent in the group with PH (32.1% vs 4.4%, p=0.001). Patients in the PH group needed a longer time of invasive ventilation than those in the no-PH group.

**Table 3:** Short-term outcomes of pulmonary hemorrhage.

| Variables | Pulmonary<br>hemorrhage<br>(n = 28) | No Pulmonary<br>hemorrhage<br>(n = 90) | p-value |
| --- | --- | --- | --- |
| Death, n (%)<sup>*</sup> | 18 (64.3) | 23 (25.6) | 0.001 |
| Intraventricular hemorrhage grade II-IV, n (%)<sup>†</sup> | 9 (32.1) | 4 (4.4) | 0.001 |
| Moderate- severe BPD, n (%)<sup>†</sup> | 5 (17.9) | 11 (12.2) | 0.528 |
| Stage 3 ROP, n (%)<sup>†</sup> | 2 (7.1) | 7 (7.8) | 1.00 |
| Length of invasive ventilation (days)<sup>‡</sup> |  |  |  |
| IMV | 5.4 (1.12; 14.4) | 0.5 (0; 4) | 0.001 |
| HFO | 1.75 (1; 6.75) | 0 (0; 1.56) | 0.001 |
| Length of using NCPAP (days)<sup>‡</sup> | 2 (0; 25) | 11 (3; 25) | 0.016 |
| NICU stay (days)<sup>‡</sup> | 21 (3.75; 52) | 24 (7; 50.25) | 0.604 |

<sup>\*</sup>Chi-square test, <sup>†</sup>Fisher's test, <sup>‡</sup> Mann-Whitney test.
Abbreviations: BPD (Bronchopulmonary Dysplasia), HFO (High-Frequency Oscillations), IMV (Intermittent Mandatory Ventilation), NICU (Neonatal Intensive Care Unit), ROP (Retinopathy of Prematurity).

## Discussion

PH is thought to occur in up to 4% to 12% of VLBW infants ^4^. Similarly, the incidence of PH was 54/1350 (4%) in a prospective cohort study by Gezmu et al., which was conducted in a tertiary-level hospital in Botswana that included 1350 newborns enrolled during a 2-year period^8^. The population of that study had a mean (SD) birth weight was 2154 (±997.5) grams and a gestational age was 34.3 (±4.7) weeks. However, the incidence of PH in our center accounts for 23.7% among newborns with gestational age at birth below 32 weeks. The mean gestational age of our patients is lower than those of Gezmu’s study (25.5 weeks vs. 34.3 weeks), and it may affect the rate of PH.

The mean age of onset of PH varied between 40 - 72 hours ^9,10^. More than 80% of cases of PH onset within 3 days of life with a median of 40 hours^10^. In 2013, a study by Yen TA et al. demonstrated that the mean age of infants who have birth weight < 1500gr with PH occurrence was 2.4 ± 1.3 days^11^. Additionally, the study of Wang et al. revealed that the median age of PH occurrence was 3 (2-4,5) days in extremely low birth weight (ELBW) infants ^3^. Similarly, the mean time for the onset of PH was 47 hours after birth in our study. In our study, we assessed risk factors for PH by multivariate analysis and identified that intubation in the first 24 hours, blood transfusion prior to PH in the first 3 days, and coagulation problems in the first 3 days were independently associated factors for PH. Similar to our result, Ferreira et al. also found that preterm infants who needed intubation in the delivery room were associated with PH occurence^4^. As can be seen in our study design, Children’s Hospital 1 has a level IV NICU, where obtaining patients’ referrals from nearby regional hospitals and intubation may be indicated conditionally for airway control while transferring. The need for intubation and positive pressure ventilation can cause excessive alveolar distension and lead to stress damage to the alveolar capillaries, which may activate the genesis of PH^12^.

In the multiple logistic regression, we found there is an association between PH and usage of blood products prior to PH in the first 3 days of life. Transfusion of blood products is thought to increase pulmonary capillary pressure, leading to stress injury of the capillary wall ^4^. Additionally, a clinical disorder which is known as transfusion-related acute lung injury (TRALI) can be detected in patients who experience a great deal of stress, which may impact the occurrence of PH^13^. TRALI begins with increasing the levels of interleukin-8 and interleukin-6, which results in neutrophil recruitment to the pulmonary vasculature, and pulmonary edema can eventually happen. The onset of TRALI can be during or from 6–72 hours after a transfusion.

Coagulation dysfunction in the first 3 days of life also showed an association with PH in a multivariate logistic regression model. In 2021, a study by Jing Li et al. found that PDA, coagulopathy, and 10-minute Apgar Score were risk factors for PH^14^. Coagulation dysfunction in preterm infants can be related to hypoxia due to respiratory distress syndrome (RDS) and sepsis. On the other hand, preterm infants may need surfactant replacement in the case of RDS, and surfactant was reported to have an impact on the impairment of coagulation in-vitro^15^.

Prior studies reported the association of PDA with the occurrence of PH^1,14^. From 2 days of age, the reduction of pulmonary vascular resistance leads to increased left-to-right shunting through PDA and pulmonary blood flow and results in elevating the pressure state of the pulmonary vessels, which may compromise cardiac function with an increased risk of PH. However, we do not reveal this association when analyzed in a multivariate logistic regression model. It can be explained that the preterm infants less than 28 weeks in our center had been given prophylactic treatment with acetaminophen and fluid restriction to reduce the risk of hsPDA.

Several studies showed that surfactant replacement may increase the risk of PH ^2,4,16^, whereas other studies reported no impact of surfactant replacement ^3,17^. In the current study, our data showed that PH was associated with surfactant replacement within 24 hours of age and repeated surfactant therapy. It is reasonable to postulate that the illness of infants who need surfactant are clinically severe and more likely to have PH than those who do not need surfactant. However, multivariate logistic regression analysis revealed no interactive effects of surfactant and PH. This is in agreement with some of the previous reports^17,18^.

For short-term outcomes of PH, we found a strong association between PH and mortality rate, similar to previous studies^2,5,17^. Another significant outcome observed in this study was IVH. PH and IVH were reported as a result of hemodynamic instability and coagulation disorders ^8^. The study by Wang TT et al. found that the IVH rate of the group of extremely low birth weight infants (ELBWIs) with PH was significantly higher than those of the group of ELBWIs without PH^3^. In our study, the rate of grade II-IV IVH was significantly higher in infants with PH than those without PH (32.1% and 4.4%, respectively, P <0.001). On the other hand, IVH can also be found in 24.4% of extremely preterm infants with hsPDA, but there has been no evidence that the presence of a hsPDA is associated with a higher probability of IVH. As mentioned above, the incidence of hsPDA was 48.3% among our patients, and it was higher in the PH group compared to the no-PH group but not statistically significant.

Supportive management of PH was positive ventilation, blood transfusion, and vasopressor administration ^19^. In our study, the duration of invasive ventilation and nCPAP support was significantly longer in the PH group than in the no-PH group (p=0.001). However, there were no significant differences in NICU stay between surviving infants in the PH and no-PH groups (p=0.6).

Our unit is a level IV NICU that primarily cares for out-born infants transferred from neonatal units in the Obstetric Hospitals in Ho Chi Minh City and the surrounding regions. This was a retrospective study in a single center, which may not be able to highlight all the associated factors of PH in very preterm newborns due to the limited data and small sample size. However, analyzing the risk factors of PH will help physicians to understand better why PH occurs and how to prevent it.

## Conclusion

PH is a severe and dreadful complication that has a common onset at 2-4 days of age in the population of newborns with <32 weeks, particularly in extremely preterm infants. The risk of PH increases with decreasing gestational age of live-born infants, especially those who have been intubated for mechanical ventilation on the first day of life, have coagulation problems within the first 3 days of life, and have blood products transfusion within the first 3 days. IVH should be investigated as soon as the diagnosis of PH is confirmed.

## Acknowledgments

We would like to acknowledge all participants who were enrolled in the study and their parents and families. We would like to acknowledge our supporting colleagues at the University of Medicine and Pharmacy at Ho Chi Minh City and Children’s Hospital 1 in Ho Chi Minh City.

## Author contributions

Conceptualization: TTN, GTMT; Data collection: GTMT, TTN; Formal analysis: GTMT, TTN, NPMN, NPL, DNN; Methodology: TTN, GTMT, NPMN, NPL, DNN; Supervision: TTN; Writing draft manuscript: GTMT, TTN, NPMN, NPL, DNN; Review and edit manuscript: GTMT, TTN, NPMN, NPL, DNN; Approve the final manuscript: all authors.

## Data availability statement

Data for this study are contained within patient medical records and are not publicly available due to privacy restrictions. The data are available from the corresponding author, Thu-Tinh Nguyen, upon reasonable request.

